# Novel prefrontal synthesis intervention improves language in children with autism

**DOI:** 10.1101/2019.12.10.19014159

**Authors:** Andrey Vyshedskiy, Edward Khokhlovich, Rita Dunn, Alexander Faisman, Jonah Elgart, Lisa Lokshina, Yuriy Gankin, Simone Ostrovsky, Lauren deTorres, Stephen M Edelson, Petr O Ilyinskii

## Abstract

Prefrontal synthesis (PFS) is defined as the ability to juxtapose mental objects at will. Paralysis of PFS may be responsible for the lack of comprehension of spatial prepositions, semantically-reversible sentences, and recursive sentences observed in 30 to 40% of individuals with ASD. In this report we present data from a three-year-long clinical trial of 6,454 ASD children aged 2-12 years, which were administered a PFS-targeting intervention. Tablet-based verbal and nonverbal exercises emphasizing mental-juxtaposition-of-objects were organized into an application called *Mental Imagery Therapy for Autism* (MITA). The test group included participants who completed more than one thousand exercises and made no more than one error per exercise. The control group included the rest of participants. The test group participants were matched to the control group by age, gender, expressive language, receptive language, sociability, cognitive awareness, and health at the 1^st^ evaluation. The test group showed 1.7-fold improvement in receptive language score vs. control group (p=0.0002) and 1.4-fold improvement in expressive language (p=0.0144). No statistically significant change was detected in other subscales not targeted by the exercises. These findings show that language acquisition improves by training PFS and warrants further investigation of the PFS-targeting intervention in a randomized controlled study.

## Introduction

Full command of complex language depends on understanding of vocabulary as well as on the mechanism of juxtaposition of mental objects into novel combinations, called Prefrontal Synthesis (PFS) ^1^. Without developed PFS it is impossible to understand the difference between sentences with identical words and grammar, such as “the cat on the mat” and “the mat on the cat.” Most people anthropomorphically assume innate PFS abilities in all individuals. Scientific evidence, however, suggests a more intricate story. While propensity toward PFS is innate in humans, acquisition of PFS seems to be the function of using recursive language in early childhood ^2–5^. The myelination of frontoposterior fiber tracts mediating PFS ^6^ depends on early childhood conversations ^7,8^. In the absence of normal recursive conversations, children do not fine-tune these neurological connections and, as a result, do not acquire PFS ^9^. The autism community refers to the phenomenon whereby individuals cannot combine disparate objects into a novel mental image as *stimulus overselectivity*, or *tunnel vision*, or *the lack of multi-cue responsivity* ^10–13^. Failure to juxtapose mental objects, called *PFS paralysis*, results in life-long inability to understand spatial prepositions, semantically-reversible sentences (e.g., “the cat on the mat”), and recursion. Among individuals diagnosed with ASD, the prevalence of PFS paralysis is 30 to 40% ^14^ and may be as high as 60% among children enrolled into special ASD schools ^15^.

We hypothesized that early PFS-targeting intervention can improve language ability in children with ASD. Accordingly, we designed various developmental activities, all of which follow a systematic approach to train PFS verbally as well as outside of the verbal domain ^16–19^. To make these activities dynamic and attractive to children, we organized them into an application called *Mental Imagery Therapy for Autism* (MITA). The MITA app was released in 2015 and quickly rose to the top of the “autism apps” charts ^16^.

MITA verbal activities start with simple vocabulary-building exercises and progress toward exercises aimed at higher forms of language, such as noun-adjective combinations, spatial prepositions, recursion, and syntax ^17^. E.g., a child can be instructed to *select the {small/large} {red/blue/green/orange} ball*, or to *put the cup {on/under/behind/in front of} the table*. All exercises are deliberately limited to as few nouns as possible since the aim is not to expand a child’s one-word vocabulary, but rather to teach him/her to integrate mental objects in novel ways by utilizing PFS ^17^.

MITA activities outside of the verbal domain aim to provide the same PFS training visually through implicit instructions as has been described in Ref. ^18^. E.g., a child can be presented with two separate images of a train and a window pattern, and a choice of complete trains. The task is to find the correct complete train and to place it into the empty square. This exercise requires not only attending to a variety of different features in both the train and its windows, but also combining two separate pieces into a single image (in other words, mentally *integrating* separate train parts into a single unified gestalt). As levels progress, the exercises increase in difficulty, requiring attention to more and more features and details. Upon attaining the most difficult levels, the child must attend to as many as eight features simultaneously. Previous results from our studies have demonstrated that children who cannot follow the explicit verbal instruction can often follow an equivalent command implicit in the visual set-up of the puzzle ^17^.

PFS is an internal, subjective function that does not easily manifest itself to evaluators ^15^. Most receptive language tests are based on vocabulary assessment and may miss the profound deficit in PFS. Furthermore, unlike vocabulary acquisition, PFS takes many years to develop in an ASD child. With long intervention time and in the absence of accepted PFS tests, randomized controlled trial (RCT) of a PFS-targeting intervention is an arduous proposition. We were not able to generate support for a long RCT and therefore resolved to conduct a simpler observational trial.

We have previously described a framework for investigating targeted interventions for ASD children epidemiologically, whereby caregivers submit multiple assessments longitudinally ^20^. When a single parent completes the same evaluation over multiple years, changes in the score become a meaningful measure of child’s progress despite a possible bias in the absolute score. Using the comprehensive 77-question Autism Treatment Evaluation Checklist (ATEC) ^21^ over the period of several years we have demonstrated significant differences in outcomes along several parameters ^20^. Younger children improved more than the older children in all four ATEC subscales – Language, Sociability, Cognitive awareness, and Health. Children with milder ASD demonstrated higher improvement in the Language subscale than children with more severe ASD. No difference between females vs. males was registered in all cohorts studied.

In this report we apply the same framework to study the PFS intervention called MITA in children aged 2 to 12 years. The data collected over five years shows greater language improvement in MITA-engaged children compared to controls matched by age, gender, expressive language, receptive language, sociability, cognitive awareness, and health at the 1^st^ evaluation.

## Methods

### MITA exercises

MITA includes both verbal and nonverbal exercises aiming to develop voluntary imagination ability ^22^ in general and Prefrontal Synthesis (PFS) ability in particular ^15^. The fidelity, validity and reliability of the MITA was discussed in detail in Refs. ^16–19^. MITA verbal activities use higher forms of language, such as noun-adjective combinations, spatial prepositions, recursion, and syntax ^17^ to train PFS: e. g., a child can be instructed to put the *large red dog behind the orange chair*, Figure 1A; or *identify the wet animal* after *the lion was showered by the monkey*; or *take animals home* following an explanation that *the lion lives above the monkey and under the cow*, Figure 1B. In every activity a child listens to a short story and then works within immersive interface to generate an answer. Correct answers are rewarded with pre-recorded encouragement and flying stars. To avoid routinization, all instructions are generated dynamically from individual words. Collectively, verbal activities have over 10 million different instructions, therefore a child will almost never hear the same instruction once again.

**Figure 1.**
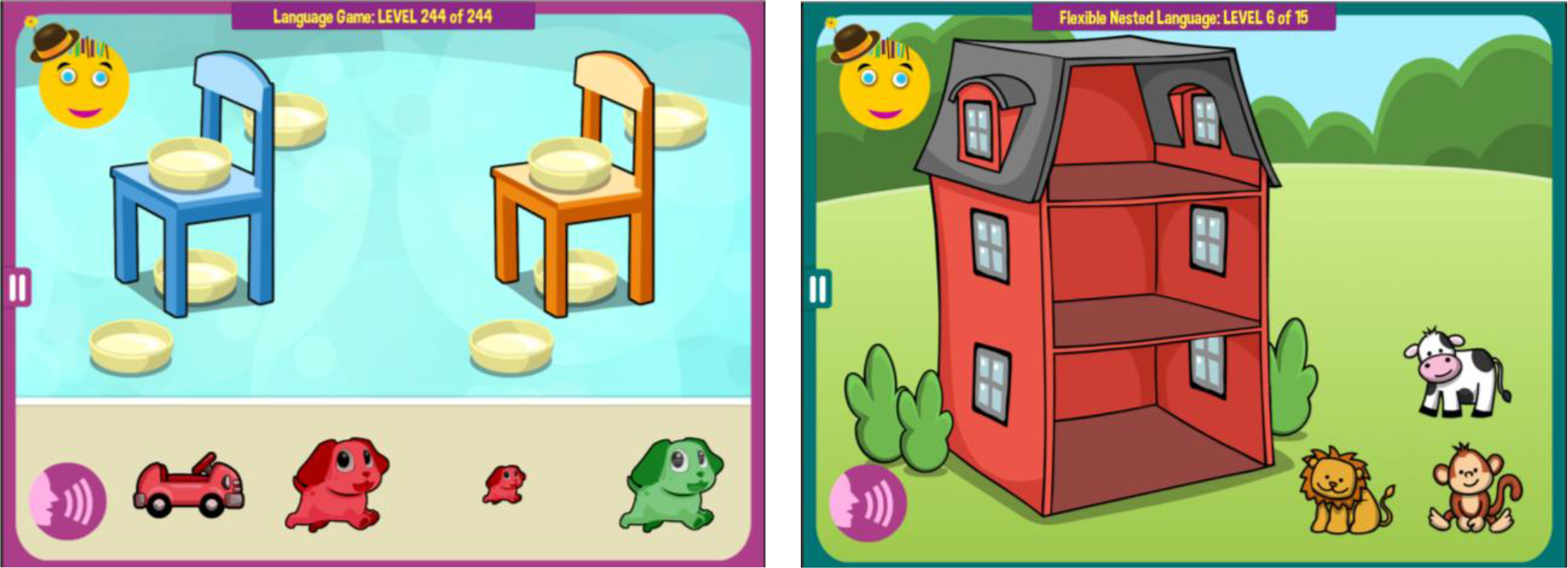
Examples of MITA verbal exercises. (A) A child is instructed to put the *large red dog behind the orange chair*. (B) A child is instructed: *Imagine. The lion lives above the monkey and under the cow. Take animals home*. Note that animals cannot be dragged to their apartments during instructions, encouraging a child to imagine animals’ correct positions in the mind.

MITA nonverbal activities aim to provide the same PFS training visually through implicit instructions ^**18**^. E.g., a child can be presented with two separate images of a train and a window pattern, and a choice of complete trains. The task is to find the correct complete train. The child is encouraged to avoid trial-and-error and integrate separate train parts mentally, thus training PFS, Figure 2A. Different games use various tasks and visual patterns to keep the child engaged, Figure 2B. Most puzzles are assembled dynamically from multiple pieces in such a way that they never repeat themselves.

**Figure 2.**
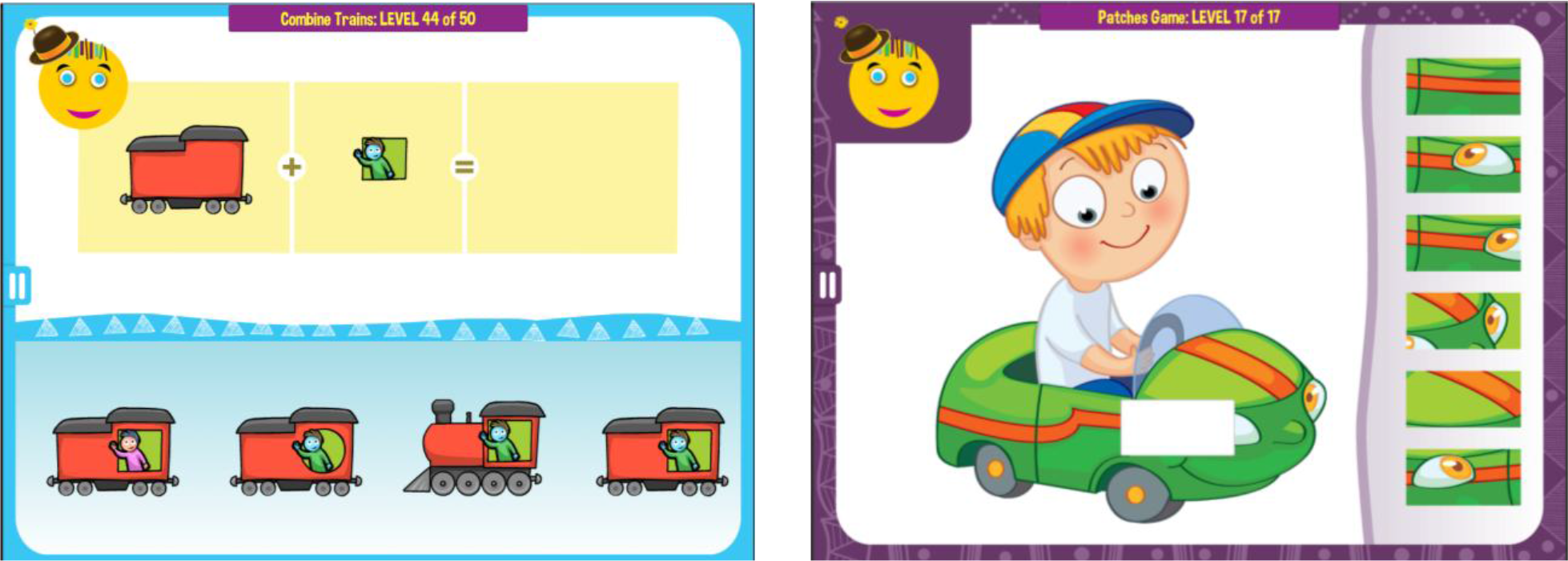
Examples of MITA nonverbal exercises. (A) Implicit instruction: *Find the correct train*. (B) Implicit instruction: *Find the correct patch*.

MITA also includes a number of hybrid activities that start children on easier nonverbal exercises and then gradually increase in difficulty, first to a combination of a verbal instruction and a visual clue and later to a verbal instruction alone, Figure 3. Collectively, MITA activities are designed to last for approximately 10 years.

**Figure 3.**
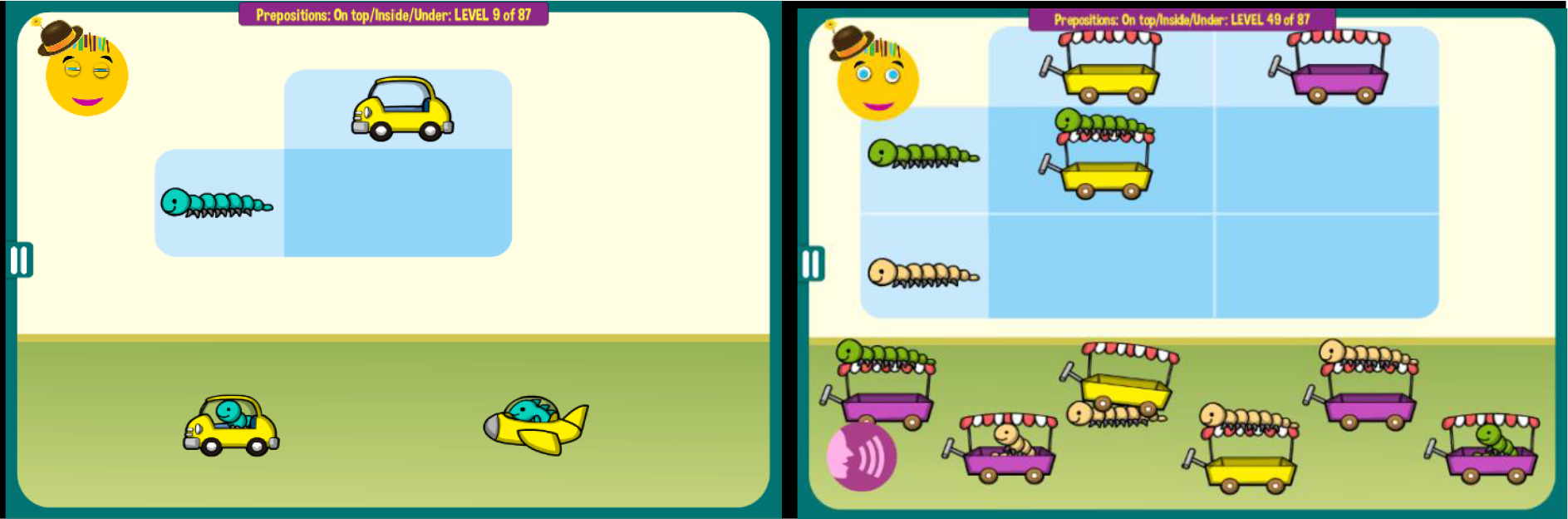
Examples of a MITA game that teach spatial prepositions *above* and *under*. (A) The game starts with the implicit instruction to combine an animal and a vehicle. (B) At more difficult levels a child must notice the correct positioning of the animal (*above* or *under*), which is announced verbally and also indicated by a visual clue in the top left corner of the matrix. At the most difficult levels (not shown) the visual clue is hidden and a child must rely on the verbal instruction alone.

### Participants

The MITA app was made available gratis at all major app stores in September 2015. Once the app was downloaded, the caregiver was asked to register and to provide demographic details, including the child’s diagnosis and age. Caregivers consented to anonymized data analysis and completed Autism Treatment Evaluation Checklist (ATEC) ^21^ as well as an evaluation of the receptive language using MSEC checklist ^23^. The first evaluation was administered approximately one month after the first use of MITA and once 100 puzzles had been completed. The subsequent evaluations were administered at approximately three-month intervals. To enforce regular evaluations, MITA app became unusable at the end of each three-month interval and parents needed to complete an evaluation to regain its functionality.

From this pool of potential study participants, we selected participants based on the following criteria:

1. **Consistency:** Participants must have filled out at least three ATEC evaluations and the interval between the first and the last evaluation was six months or longer.
2. **Diagnosis:** The subjects must have self-reported their diagnosis as ASD. The ASD diagnosis was not verified directly, as we cannot ask participants to submit documentation. However, ATEC scores support ASD diagnosis. Average initial ATEC total score in the test group was 68.71 ± 24.26, and 67.95 ± 23.68 in the control group, Table 1, which corresponds to medium-to-severe ASD as delineated in Ref. ^24^ and Table 2.
3. **Maximum age:** Participants older than twelve years of age were excluded from this study.
4. **Minimum age:** Participants who completed their first evaluation before the age of two years were excluded from this study.

**Table 1.** Test and control group characteristics at the 1^st^ evaluation. A lower score indicates a lower severity of ASD symptoms.

|  | **Test** | **Control** | **P-value** |
| --- | --- | --- | --- |
| **Participants in each age group (total)** | 1009 | 1009 | n/a |
| **Age at baseline (mean ± SD)** | 5.3 ± 2.1 | 5.3 ± 2.4 | 0.87 |
| **Male Gender** | 74% | 75% | 0.90 |
| **Receptive Language (mean ± SD)** | 14.4 ± 6.2 | 14.5 ± 6.4 | 0.26 |
| **Expressive Language (mean ± SD)** | 14.4 ± 6.2 | 14.5 ± 6.4 | 0.21 |
| **Sociability (mean ± SD)** | 14.6 ± 7.0 | 14.7 ± 7.1 | 0.67 |
| **Cognitive Awareness (mean ± SD)** | 16.4 ± 6.3 | 16.2 ± 6.7 | 0.91 |
| **Health (mean ± SD)** | 21.5 ± 11.4 | 22.0 ± 12.3 | 0.07 |
| **ATEC Total (mean ± SD)** | 68.71 ± 24.26 | 67.95 ± 23.68 | 0.89 |

**Table 2.** Approximate relationship between ATEC total score, age, and ASD severity as described elsewhere ^20^

|  | Age |  |  |  |  |  |  |  |  |  |  |
| --- | --- | --- | --- | --- | --- | --- | --- | --- | --- | --- | --- |
| Severity | 2 | 3 | 4 | 5 | 6 | 7 | 8 | 9 | 10 | 11 | 12 |
| Mild | <82 | <65 | <52 | <43 | <36 | <31 | <28 | <25 | <23 | <21 | <20 |
| Moderate | 82-130 | 65-103 | 52-83 | 43-69 | 36-58 | 31-50 | 28-44 | 25-39 | 23-36 | 21-34 | 20-32 |
| Severe | 130-179 | 103-179 | 83-179 | 69-179 | 58-179 | 50-179 | 44-179 | 39-179 | 36-179 | 34-179 | 32-179 |

After excluding participants that did not meet these criteria, there were 6,454 total participants, from whom we have selected the test and control groups, Table 1.

### Test and control groups

Since our application was available for free to the general public, there was a large volume of downloads by people of widely ranging commitment. To assess the effect of MITA intervention we had to identify participants who have not just downloaded MITA and used trial-and-error to arrive at solutions, but actively engaged with MITA. Thus, we have selected participants who completed at least one thousand exercises and made no more than one error per exercise (N=1,009). The test group participants were matched to the control group by age, gender, expressive language, receptive language, sociability, cognitive awareness, and health at 1^st^ evaluation (baseline) using propensity score analysis ^25^.

### Outcome measures

A caregiver-completed Autism Treatment Evaluation Checklist (ATEC) ^21^ and Mental Synthesis Evaluation Checklist (MSEC) ^23^ were used to track the efficacy of the treatment. The complete ATEC questionnaire (can be accessed freely at www.autism.org) is comprised of four subscales: 1) Speech/Language/Communication, 2) Sociability, 3) Sensory/Cognitive Awareness, and 4) Physical/Health/Behavior. The first subscale, Speech/Language/Communication, contains 14 items and its score ranges from 0 to 28 points. The Sociability subscale contains 20 items within a score range from 0 to 40 points. The third subscale, referred here as the Cognitive Awareness subscale, has 18 items and scores range from 0 to 36 points. The fourth subscale, referred here as the Health subscale contains 25 items and scores range from 0 to 75 points. The scores from each subscale are combined in order to calculate a Total Score, which ranges from 0 to 179 points. A lower score indicates lower severity of ASD symptoms and a higher score indicates more severe symptoms of ASD. ATEC is not a diagnostic checklist. It was designed to evaluate treatment effectiveness ^21^ and ASD severity can be related to ATEC total score and age only approximately. Table 2 lists approximate ATEC total score as related to ASD severity and age as described elsewhere ^20^.

ATEC was selected because it is one of the few measures validated to evaluate treatment effectiveness. In contrast, another popular ASD assessment tool, ADOS, ^26^ has been only validated as a diagnostic tool. Various studies confirmed validity and reliability of ATEC ^27–29^ and several trials confirmed ATEC’s ability to longitudinally measure changes in participant performance ^20,30–32^. Whitehouse et al. used ATEC as a primary outcome measure for a randomized controlled trial of their iPad-based intervention for ASD named TOBY and noted ATEC’s “internal consistency and adequate predictive validity” ^33^. These studies support the effectiveness of ATEC as a tool for longitudinal tracking of symptoms and assessing changes in ASD severity.

### Expressive language assessment

ATEC Speech/Language/Communication subscale starts by assessing the simplest linguistic abilities, such as 1) Knows own name, 2) Responds to ‘No’ or ‘Stop’, 3) Can follow some commands, 4) Can use one word at a time (No!, Eat, Water, etc.), 5) Can use 2 words at a time (Don’t want, Go home), 6) Can use 3 words at a time (Want more milk) and progress to interrogate complex language abilities, such as 7) Knows 10 or more words, 8) Can use sentences with 4 or more words, 9) Explains what he/she wants, 10) Asks meaningful questions, 11) Speech tends to be meaningful/relevant, 12) Often uses several successive sentences, 13) Carries on fairly good conversation, and 14) Has normal ability to communicate for his/her age. With the exception of the first three items, all the Language subscale items primarily depend on expressive language. Accordingly, the ATEC subscale 1 is referred in this manuscript as Expressive Language subscale to distinguish it from the Receptive Language subscale tested by the MSEC evaluation.

### Receptive language assessment

MSEC evaluation was designed to be complementary to ATEC in measuring receptive language and PFS. Out of 20 MSEC items those that directly assess receptive language are the following: 1) Understands simple stories that are read aloud; 2) Understands elaborate fairy tales that are read aloud (i.e. stories describing FANTASY creatures); 6) Understands some simple modifiers (i.e. green apple vs. red apple or big apple vs. small apple); 7) Understands several modifiers in a sentence (i.e. small green apple); 8) Understands size (can select the largest/smallest object out of a collection of objects); 9) Understands possessive pronouns (i.e. your apple vs. her apple); 10) Understands spatial prepositions (i.e. put the apple ON TOP of the box vs. INSIDE the box vs. BEHIND the box); 11) Understands verb tenses (i.e. I will eat an apple vs. I ate an apple); 12) Understands the change in meaning when the order of words is changed (i.e. understands the difference between ‘a cat ate a mouse’ vs. ‘a mouse ate a cat’); 20) Understands explanations about people, objects or situations beyond the immediate surroundings (e.g., “Mom is walking the dog,” “The snow has turned to water”). MSEC consists of 20 questions and is scored similarly to ATEC: a lower score indicates better receptive language and PFS ability. To simplify interpretation of figure labels, the subscale 1 of the ATEC evaluation is referred to as the Expressive Language subscale and the MSEC scale is referred as the Receptive Language subscale.

### Statistical analysis

The framework for evaluation of ATEC score changes over time was explained in detail earlier ^20^. In short, the concept of a “Visit” was developed by dividing the three-year-long observation interval into 3-month periods. All evaluations were mapped into 3-month-long bins with the first evaluation placed in the first bin. When more than one evaluation was completed within a bin, their results were averaged to calculate a single number representing this 3-month interval. It was then hypothesized that there was a two-way interaction between Visit and treatment. Statistically, this hypothesis was modeled by applying the Linear Mixed Effect Model with Repeated Measures (MMRM), where a two-way interaction term was introduced to test the hypothesis. The model (Endpoint ∼ Baseline + Gender + Severity + Treatment * Visit) was fit using R Bioconductor library of statistical packages, in particular “nlme” package. The subscale score at baseline, gender, and severity were used as covariates. Conceptually, the model fits a plane into n-dimensional space. This plane takes into account a complex variability structure across multiple visits, including baseline differences. Once such plane is fit, the model calculates Least Squares Means (LS Means) for each subscale and treatment group at each visit. The model also calculates LS Mean differences between the treatment and control groups at each visit. Participants in the test group were matched to those in the control group using propensity score analysis ^25^ based on age, gender, expressive language, receptive language, sociability, cognitive awareness, and health at the 1^st^ evaluation (baseline).

### Informed Consent

Caregivers have consented to anonymized data analysis and publication of the results.

### Clinical Trial Registration

The observational clinical trial, ClinicalTrials.gov Identifier: NCT02708290, was registered on March 15, 2016.

### Compliance with Ethical Standards

Using the Department of Health and Human Services regulations found at 45 CFR 46.101(b)(1), it was determined that this research project is exempt from IRB oversight.

### Data Availability

De-identified raw data from this manuscript are available from the corresponding author upon reasonable request.

### Code availability statement

Code is available from the corresponding author upon reasonable request.

## Results

We first sought to replicate our earlier results ^20^ using the new database. The analysis of groups within the MITA database was consistent with our previous analysis performed on the database collected by the Autism Research Institute. There was no difference between females vs. males in any subscale. Younger children improved more than the older children in the Language subscale (Tables S1, S2). Children with milder ASD improved more than children with more severe ASD in the Language subscale (Tables S3, S4).

Having demonstrated continuity with respect to group differences within MITA database, we have applied the same statistical framework to study the difference between the test group and the control group. Children in both the control and test groups improved their symptoms over time in all subscales. The greatest interest was the Receptive Language subscale targeted by the MITA intervention. The average improvement in test group over three years was 8.49 points (SE=0.65, p<0.0001) compared to 4.87 points (SE=0.85, p<0.0001) in the control group, Figure 4A, Table 3. The difference in the test group relative to the control group at Visit 12 was statistically significant: −3.93 points (SE=1.05, p=0.002). The negative (test-control) indicates that the test group had lower score at visit 12 and therefore milder symptoms. On the annualized basis, the test group improved their receptive language 1.7-times faster than the control group (Test=2.8 points/year; Control=1.6 points/year).

**Table 3:** LS Means for test and control groups. Data are presented as: LS Mean (SE; 95% CI). The differences between Test and Control and between Visit 12 and Visit 1 are presented as LS Mean (SE; P-value). A lower score indicates a lower severity of ASD symptoms. The negative Test-Control indicates that the Test group had lower score at visit 12 and therefore milder symptoms.

|  | Visit 1 |  |  | Visit 12 |  |  | Visit 12 - Visit 1 |  |
| --- | --- | --- | --- | --- | --- | --- | --- | --- |
|  | Test | Control | Test - Control | Test | Control | Test - Control | Test | Control |
| **Receptive Language** | 26.7 (0.27; 26.2 - 27.2) | 27 (0.27; 26.5 - 27.5) | -0.31 (0.28; 0.2608) | 18.2 (0.66; 16.9 - 19.5) | 22.1 (0.86; 20.5 - 23.8) | -3.93 (1.05; 0.0002) | -8.49 (0.65; <0.0001) | -4.87 (0.85; <0.0001) |
| **Expressive Language** | 13.89 (0.2; 13.51 - 14.28) | 14.14 (0.19; 13.76 - 14.51) | -0.24 (0.19; 0.2141) | 8.87 (0.45; 7.98 - 9.75) | 10.6 (0.58; 9.46 - 11.74) | -1.74 (0.71; 0.0144) | -5.03 (0.43; <0.0001) | -3.53 (0.57; <0.0001) |
| **Sociability** | 13.7 (0.25; 13.2 - 14.2) | 13.8 (0.25; 13.3 - 14.3) | -0.11 (0.25; 0.6679) | 11.8 (0.61; 10.6 - 13) | 13.1 (0.79; 11.6 - 14.7) | -1.27 (0.97; 0.1875) | -1.88 (0.59; 0.0015) | -0.72 (0.78; 0.3565) |
| **Cognitive Awareness** | 15.4 (0.23; 15 - 15.9) | 15.4 (0.22; 15 - 15.8) | 0.02 (0.22; 0.9118) | 13 (0.54; 11.9 - 14) | 13.2 (0.7; 11.8 - 14.6) | -0.22 (0.85; 0.7987) | -2.43 (0.52; <0.0001) | -2.19 (0.68; 0.0014) |
| **Health** | 20 (0.39; 19.3 - 20.8) | 20.8 (0.39; 20 - 21.5) | -0.73 (0.4; 0.0678) | 18.4 (0.96; 16.5 - 20.2) | 19.4 (1.24; 16.9 - 21.8) | -1.02 (1.52; 0.5019) | -1.68 (0.94; 0.0726) | -1.38 (1.23; 0.2585) |

**Figure 4.**
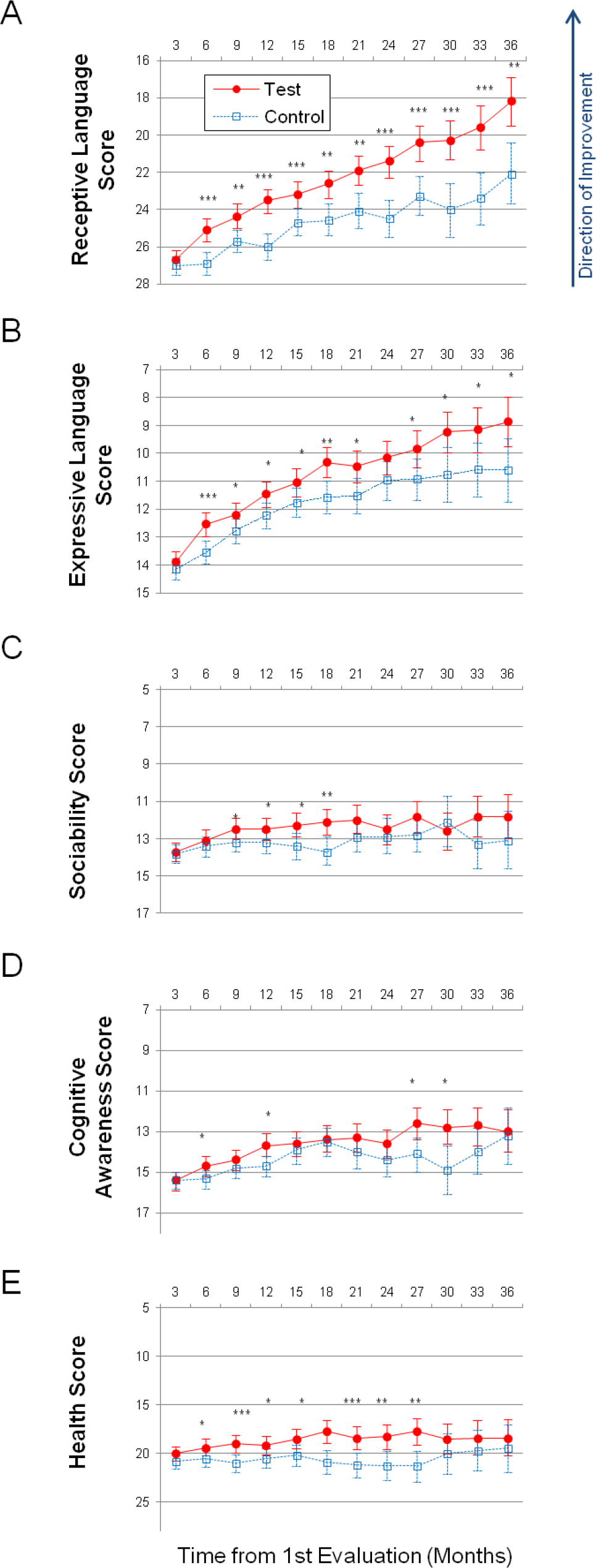
Longitudinal plots of subscale scores LS Means. Horizontal axis shows months from the 1st evaluation (0 to 36 months). Error bars show the 95% confidence interval. To facilitate comparison between subscales, all vertical axes ranges have been normalized to show 35% of their corresponding subscale’ maximum available score. A lower score indicates symptoms improvement. P-value is marked: ***<0.0001; **<0.001; *<0.05. (A) Receptive Language score. (B) Expressive Language score. (C) Sociability score. (D) Cognitive awareness score (E) Health score.

On the Expressive Language subscale, test group improved over the three-year period by 5.03 points (SE=0.43, p<0.0001) compared to 3.53 points (SE=0.57, <0.0001) improvement in the control group, Figure 4B. The difference in the test group relative to the control group at Visit 12 was statistically significant: −1.74 points (SE=0.71, p=0.014). On the annualized basis, the test group improved their expressive language 1.4-times faster than the control group (Test=1.7 points/year; Control=1.2 points/year).

On the Sociability subscale, test group improved over the three-year period by 1.88 points (SE=0.59, p=0.0015) compared to 0.72 points (SE=0.78, p=0.3565) improvement in the control group, Figure 4C. The difference in the test group relative to the control group at Visit 12 was not statistically significant.

On the Cognitive awareness subscale, test group improved over the three-year period by 2.43 points (SE=0.52, p<0.0001) compared to 2.19 points (SE=0.68, p=0.0014) in the control group, Figure 4D. The difference in the test group relative to the control group at Visit 12 was not statistically significant.

On the Health subscale, test group improved over the three-year period by 1.68 points (SE=0.94, p=0.0726) compared to control group improved 1.38 points (SE=1.23, p=0.2585), Figure 4E. The difference in the test group relative to the control group at Visit 12 was not statistically significant.

## Discussion

In this report, we described data from three-year-long clinical trial of PFS-targeting intervention – *Mental Imagery Therapy for Autism* or MITA ^16^ – that included 6,454 children with ASD. This is the longest-running and the largest study of a caregiver-administered early intervention tool for young children with ASD. Both the test and control groups improved their symptoms over time in all subscales. Control participants demonstrated similar improvement to test participants in all subscales except on the Receptive Language and the Expressive Language subscales. Test participants showed 1.7-fold greater improvement in Receptive Language score at the end of the trial compared to the control group, Figure 4A.

### Language improvement mechanisms

There are three possible explanations for greater improvement of receptive language in MITA-engaged children: 1) selection-bias of children or parents during trial, 2) indirect effect of MITA exercises through educating parents in the techniques of language therapy, and 3) direct effect of MITA exercises on neural networks essential for language. We discuss all three possibilities in detail below.

1. **Selection-bias**. Difference in language improvement between the test and control groups could be explained by selection-bias of children or parents over the course of the trial. If smarter/healthier children and more motivated parents were to continue working with MITA and remain in the trial, and weaker children/less motivated parents were to drop out from the trial, then test group would have demonstrated better outcome compared to control. However, if the smarter/healthier children and more motivated parents were self-selecting to continue the trial, one would expect to see improved outcomes in all areas of children development. On the contrary, the test group showed better outcome only in the Receptive Language subscale that was trained by MITA (test-control = −3.93, p=0.0002) and in the related Expressive Language subscale (test-control = −1.74, p=0.0144), Table 3 (the negative difference between test and control indicates that the test group had lower score at visit 12 and therefore milder symptoms). The test group outcomes in all other subscales were similar to the control group, Table 3: Sociability (test-control = −1.27, p=0.1875), Cognitive awareness (test-control = −0.22, p=0.7987), and Health subscales (test-control = −1.02, p=0.5019). Thus, selection-bias of the test group for healthier and smarter children or more motivated parents was not likely to be responsible for improvements in the Receptive Language score alone.
2. **Indirect effect of MITA exercises through educating parents in the techniques of language therapy**. Some caregivers could learn language therapy techniques used in MITA directly from MITA activities or from the “Tip of the Day.” For example, parents could learn to administer directions with increasing complexity, such as: *Give me the small woodchip* -> *Give me the small white woodchip* -> *Give me two small white woodchips -*> *Put the small white woodchip on/under the table*, etc. and then extend these techniques to everyday activities multiplying the effect of MITA exercises many-fold. A search of the MITA listing at the app store yielded several unsolicited MITA parents’ reviews, such as this one – “MITA… helps me to grab ideas from the screen and into everyday” – that supported the parents learning hypothesis. To study parents learning hypothesis further, we have solicited feedback from MITA caregivers. More than half of responders reported that they have learned language therapy techniques from MITA exercises (unpublished observations). Moreover, literature search revealed that even short-term parents’ interventions administered early have been shown to have significant effects ^34,35^ on children language acquisition, supporting the parents-learning-from-MITA-exercises hypothesis.
3. **Direct effect of MITA exercises**. It is possible that MITA exercises directly trained neural networks essential for language. Association of Wernicke’s and Broca’s areas with language is well-known. Less common is the realization that understanding of full language depends on the lateral prefrontal cortex (LPFC). Wernicke’s area primarily links words with objects ^36^, Broca’s area interprets the grammar and assigns words in a sentence to a grammatical group such as noun, verb, or preposition ^36^, but only the LPFC can synthesize the objects from memory into a novel mental image according to grammatically imposed rules ^9,37^. This latter function may be called *imagination*, but we prefer a more specific term, *Prefrontal Synthesis* (PFS) in order to distinguish this function from other components of imagination, such as dreaming, simple memory recall, spontaneous insight, mental rotation, and integration of modifiers ^1^. PFS is defined as voluntary juxtaposition of mental objects. All MITA exercises were designed to train child’s ability to combine mental objects.

PFS ability is essential for understanding sentences describing novel combinations of objects. E.g., the semantically-reversible sentences “The dog bit my friend” and “My friend bit the dog” use identical words and grammar. Appreciating the misfortune of the first sentence and the humor of the second sentence depends on the LPFC ability to faithfully synthesize the two objects – the friend and the dog – into a novel mental image. Similarly, understanding of spatial prepositions such as *in, on, under, over, beside, in front of, behind* requires a subject to synthesize several objects in front of the mind’s eye. For example, the request “to put a green box {inside/behind/on top of} the blue box” requires an initial mental simulation of the scene, only after which is it possible to correctly arrange the physical objects. An inability to produce a novel mental image of the green box {inside/behind/on top of} the blue box would lead to the use of trial-and-error, which in majority of cases will result in an incorrect arrangement.

PFS is mediated by the LPFC ^38–43^. It was hypothesized that to conduct PFS, the LPFC synchronizes object-encoding neuronal ensembles (objectNEs) in the posterior cortex (temporal, parietal, and occipital cortices) using the frontoposterior connections, such as arcuate fasciculus and superior longitudinal fasciculus (the Neuronal Ensembles Synchronization hypothesis or NES) ^1,44,45^. Patients with damage to any component of this circuit—the LPFC^39^, or the frontoposterior fibers ^6^, or temporal-parietal-occipital junction ^46^—often lose access to the full extent of PFS. Fuster calls their condition “prefrontal aphasia” ^42^ and Luria “semantic aphasia” ^47^. Fuster explains that “although the pronunciation of words and sentences remains intact, language is impoverished and shows an apparent diminution of the capacity to ‘prepositionize.’ The length and complexity of sentences are reduced. There is a dearth of dependent clauses and, more generally, an underutilization of what Chomsky characterizes as the potential for recursiveness of language.” (We prefer to call this condition ‘PFS paralysis’ since aphasia is translated from Greek as “speechless” and these patients may not experience any speech deficit.)

PFS paralysis is a known problem in individuals with ASD, commonly described as *stimulus overselectivity*, or *tunnel vision*, or *the lack of multi-cue responsivity* ^11–13^. Many techniques used by speech language pathologists (SLP) and Applied Behavioral Analysis (ABA) therapists happen to aim at improving PFS. SLPs commonly refer to these techniques as “combining adjectives, location/orientation, color, and size with nouns,” “following directions with increasing complexity,” and “building the multiple features/clauses in the sentence” ^48^. In ABA jargon, these techniques are known as “visual-visual and auditory-visual conditional discrimination” ^49–52^, “development of multi-cue responsivity” ^11^, and “reduction of stimulus overselectivity” ^12^. In some sense, MITA exercises can be viewed as an extreme version of language therapy with minimum vocabulary training and focusing nearly exclusively on mental integration techniques. MITA exercises can also be viewed as an extension of the “matrix training” ^53^ with minimal number of words and maximum number of word combinations. We conclude that the direct effect of MITA PFS exercises on language improvement cannot be excluded and in fact could be the most parsimonious explanation for the observed results.

### Nonverbal imagination exercises

The idea to use nonverbal PFS exercises in children with developmental delay dates back to Piaget, Vygotsky, and Luria ^54^. In his famous twin study, Luria used educational games developed from a set of blocks to try to improve one twin’s PFS ability. Using blocks, he designed two types of learning activities: 1) build-from-elements, and 2) build-from-model. The ‘build-from-elements’ activity involved building a structure per design that indicated contours of individual blocks necessary for construction. The ‘build-from-model’ activity indicated the overall outline of the final structure but didn’t specify what blocks to use (i.e. the design did not have contours of individual blocks indicated on it). Luria then studied ten children (5 pairs of identical twins; age 5 years). One twin of each pair followed a 2.5-month program using strategy #1 and the other using strategy #2. When tested at the end of the program, both twins in a pair were equally good at discriminating elementary figures and concentrating, but the twin following program #2 was superior in both PFS and language: he planned more, had a better sense of the relation of a block to the whole structure; program #2 twins were also *more articulate* when identifying differences between their structure and the model they were working towards ^54^.

The use of tablet computers significantly enhances the range and adaptability of nonverbal exercises. MITA nonverbal exercises are limitless in variations, therefore avoiding routinization. Each activity is dynamic, quickly adjusting to the child’s exact ability level. All activities are disguised as games that engage children. Furthermore, many MITA verbal modules start with nonverbal levels. E.g., the *Prepositions on/under* and the *Prepositions in front/behind* games have each 30 introductory nonverbal levels, whereby objects arrangement is guided by picture-example alone; 30 intermediate levels, where objects arrangement is guided by both picture-example and verbal instruction; and 30 advanced levels, where objects arrangement is guided by verbal instruction alone. Since nonverbal exercises are easier to children with ASD ^17^, MITA starts with nonverbal exercises and adds up verbal exercises slowly in order to keep the fine balance between being engaging and challenging.

### Potential issues associated with use of tablet computers for therapy administration

Giving caregivers an opportunity to administer language therapy on a tablet device comes with important warnings associated with the use of a tablet. Most apps are highly addictive to children ^55^. Many children left to their own devices will watch YouTube for hours. A tablet device introduces an opportunity for a caregiver to leave a child in what is perceived to be a safe and productive environment. A therapeutic app carries a significant danger of being a gateway to YouTube. Having said that, we note that tablet computers are already a common staple of most families, particularly families with minimally-verbal ASD children who use tablets for communication ^56^. Moreover, cognitively challenging apps like MITA are significantly less addictive, as every step requires a mental effort. Once children run out of “cognitive energy” they lose their interest in MITA. Furthermore, MITA was designed to provide minimum sensory stimulation to reduce distractions and further lower its potential for addictiveness. It is also important to stress that every parent using MITA has signed a consent form that informed them of the danger of screen time for children and explained how to lock their tablet on the MITA application. Finally, in the future, MITA can be provided on a dedicated standalone device with no ability to download other apps to completely avoid the danger of exposure to YouTube and other addictive apps.

### Why was the MITA effect on Language so significant?

Test group children showed 1.7-fold improvement in the Receptive Language score at the end of the trial vs. the control group. This result is significantly better than reported in other trials ^35,57^. There could be several reasons for greater improvement reported herein:

1. There may exist a synergy between MITA exercises and language therapy administered by SLPs and ABA therapists. SLP and ABA techniques also aim to improve PFS, but PFS exercises are just a small part of language therapy that primarily focuses on building up the child’s vocabulary. Word comprehension is a low hanging fruit. It is easier to train and also highly appreciated by parents. Furthermore, most tests rely exclusively on a child’s vocabulary to measure educational success (e.g., Peabody Picture Vocabulary Test (PPVT-4) ^58^, Expressive Vocabulary Test (EVT-2) ^59^), thus encouraging therapists to focus on vocabulary training. However, such training by itself does not train PFS that is essential for understanding of spatial preposition, recursion, and complex language. Thus, the success of MITA intervention may derive from its exclusive focus on voluntary imagination and its most advanced component, PFS.
2. Even when therapists administer PFS exercises, the training is mostly verbal in nature. The intuitive verbal approach is working well in neurotypical children, but can be abstruse for nonverbal and minimally-verbal children with ASD. MITA, on the other hand, starts with nonverbal exercises that are much easier for children with ASD ^17^.
3. Another hidden advantage of computerized language therapy for children with ASD could be its prosody stability. All MITA instructions are drawn from a pre-recorded library. Unlike human-given instructions, MITA verbal instructions are always pronounced with the same intonation. There are no variations in prosody. This prosody stability simplifies instruction interpretation for children who have auditory processing problems.
4. The most significant challenge of conventional therapy is a substantial cost that significantly reduces therapists availability to most families ^60^. The free MITA application, on the other hand, is always available and its everyday use does not increase the financial burden.
5. Unlike a human therapist, MITA is readily available for download and use within minutes. As a result, parents can initiate MITA exercises when ASD diagnosis is suspected but has not been confirmed by a clinician. In fact, many MITA parents indicate their diagnosis as “suspected ASD” at registration and change their diagnosis to ASD months later suggesting that they have started administering MITA before receiving a diagnosis from a clinician. Earlier intervention may be providing an important head-start to children.
6. To receive optimal therapy, children have to develop a certain degree of connection with a therapist ^61^. In a field in which frequent rotation of therapists is a norm, a lot of time is wasted on the initial therapist-child bridge-building ^62^. MITA, on the other hand, has only a minimal break-in period.
7. Children often miss a scheduled therapy session due to travel, sickness, tantrums, or lack of focus ^63^. Once a session has been missed, a therapist may not be available for a make-up lesson. One of the benefits of MITA is that parents can administer MITA anytime when they feel that children are in a good mood and receptive to therapy.
8. When parents delegate all therapy to professionals, they may not participate in therapy sessions, and, as a result, may never learn language therapy techniques. Consequently, the most valuable time that could have been used for parent-child communication is missed. MITA inadvertently forces parents to work with their children through language exercises, therefore promoting parents’ learning of language therapy techniques.
9. Finally, MITA gives parents hope and it is this hope that helps parents to motivate their children and persist with language therapy. By seeing their children solving puzzles, parents become more confident of their children. As one parent wrote in an unsolicited review: “My son displays an intellectual capability, which I thought for a long time was missing.”

### Limitations

The observational design of this study cannot definitively prove causality since unknown confounders may influence the study results. The golden standard of testing a novel clinical intervention is a randomized controlled trial (RCT). Prior to conducting the MITA study, we have submitted the proposal for a therapist-administered RCT of the PFS intervention to many potential funding agencies. The proposal has failed to find any traction. We have also considered a caregiver-administered RCT, but decided against it due to high attrition rate. The only published RCT of caregiver-administered tablet-based therapy for young children with ASD reported an overwhelming drop just after 3 months despite biweekly telephone calls to encourage app use ^33^. Specifically, during the first 3-month period, participants exercised for a total median time of 1,593 minutes (just under the recommended target of 20 min/day or 1,800 min/3-month period) and during the second 3-month period, participants exercised for a total median time of 23 minutes (98.6% drop in app use). In effect, most participants did not receive any intervention after the first 3-month period and therefore were lost for the RCT ^33^. As the minimal length of a PFS intervention RCT is likely to exceed two years ^57,64^, participant dropout becomes the major issue. This high attrition rate introduces multiple selection biases that devalue RCT ability to demonstrate causality and essentially makes it no better than an observational trial. The reported self-funded observational trial is the best study we could conduct without external funding.

Another disadvantage of the low-cost geographically diverse observational trials is their reliance on parent-reported outcome measures. There is an understanding within the psychological community that parents cannot be trusted with an evaluation of their own children. In fact, parents often yield to wishful thinking and overestimate their children’s abilities on a single assessment ^65^. However, the pattern of changes can be generated by measuring the score dynamics over multiple assessments. When a single parent completes the same evaluation every three months over multiple years, changes in the score become meaningful. In this trial evaluations were administered at regular 3-month intervals.

Our results should be treated cautiously, as less motivated families may not be able to commit themselves to long-term therapy administration and families without technical backgrounds may find their experience with MITA less intuitive. Further validation of MITA exercises is necessary to understand its efficacy within the diverse ASD population.

## Conclusions

Three major conclusions follow up from this study. First, Prefrontal Synthesis (PFS) is an essential component of full language and exercises training PFS are an indispensable component of language therapy. Second, some caregivers are capable of administering tablet-based exercises to their children consistently over many years. Third, parent-administered and parent-reported multiyear observational trials can be an attractive low-cost model for studying novel language, behavioral, and dietary interventions. The significant improvement of receptive language observed in the current trial brings hope to many families and inspires us to continue developing PFS exercises and to translate MITA to multiple languages. The major strength of this study is the large number of long-term participants. The most obvious limitation is that this study observational design cannot definitively prove causality since not all confounders can be adjusted appropriately. We conclude that the MITA PFS intervention warrants further investigation in a randomized controlled study (RCT).

## Acknowledgments

We wish to thank Dr. Noah Greifer for assistance with statistical analysis, Danielle Abate for assistance with manuscript preparation and Yulia Dumov for the design of children’s activities.

## Author contributions

AV, EK, and POI designed the study. RD, AF, JE, LL, YG and AV developed the MITA app. SME acquired the treatment as usual data. EK, SO, LdT, and AV analyzed the data. AV, EK, and POI wrote the paper.

**Supplementary Information** is linked to the online version of the paper.

