## Supplemental Material for "Novel prefrontal synthesis intervention improves language in children with autism"

Tables S1 to S4 replicate our previously published analysis (Mahapatra, Shreyas, et al. "Longitudinal Epidemiological Study of Autism Subgroups Using Autism Treatment Evaluation Checklist (ATEC) Score." *Journal of autism and developmental disorders* (2018): 1-12) on the MITA database. Since the previous analysis was conducted over the two-year-period, Tables S1 to S4 only report LS Mean differences at the baseline (Visit 1) and at the end of the two-year period (Visit 8). The rest of the tables report the analysis of the three-year period from Visit 1 to Visit 12.

**Table S1. MITA participants database: LS Mean differences between Visit 8 and Visit 1. Data are presented as LS Mean (SE; P-value).**

|  | **Visit 8 – Visit 1** | | |
| --- | --- | --- | --- |
|  | 2-3 YOA | 3-6 YOA | 6-12 YOA |
| **Subscale 1: Language** | -7.59 (0.35; <0.0001) | -4.23 (0.21; <0.0001) | -1.24 (0.39; 0.47) |
| **Subscale 2: Sociability** | -2.23 (0.48; <0.004) | -2.30 (0.29; <0.0001) | -1.43 (0.54; 0.88) |
| **Subscale 3: Cognitive**  **Awareness** | -2.8 (0.41; <0.0001) | -1.60 (0.24; <0.0001) | -0.17 (0.47; 1.00) |
| **Subscale 4: Health** | 0.08 (0.76; 1.00) | -1.75 (0.46; 0.08) | -0.62 (0.86; 1.00) |

**Table S2. MITA participants database: LS Mean differences between Age Groups. Data are presented as: LS Mean difference (SE; P-Value)**

|  | **Visit 1** | | | **Visit 8** | | |
| --- | --- | --- | --- | --- | --- | --- |
|  | 2-3 vs. 3-6 | 2-3 vs. 6-12 | 3-6 vs. 6-12 | 2-3 vs. 3-6 | 2-3 vs. 6-12 | 3-6 vs. 6-12 |
| **Subscale 1: Language** | 1.14 (0.20; <0.0001) | 1.61 (0.28; <0.0001) | 0.47 (0.22; 0.99) | -2.22 (0.39; <0.0001) | -4.73 (0.52; <0.0001) | -2.51 (0.43; <0.0001) |
| **Subscale 2: Sociability** | 0.75 (0.26; <0.72) | 1.10 (0.35; <0.55) | 0.35 (0.28; 1.00) | 0.82 (0.53; 1.00) | 0.29 (0.70; 1.00) | -0.52 (0.58; 1.00) |
| **Subscale 3: Cognitive**  **Awareness** | 0.75 (0.22; 0.32) | 0.86 (0.31; 0.76) | 0.11 (0.24; 1.00) | -0.46 (0.46; 1.00) | -1.77 (0.60; 0.66) | -1.32 (0.50; 0.89) |
| **Subscale 4: Health** | 0.40 (0.42; 1.00) | 0.78 (0.58; 1.00) | 0.38 (0.46; 1.00) | 2.23 (0.85; 0.88) | 1.48 (1.12; 1.00) | --0.75 (0.94; 1.00) |

**Table S3. MITA participants database, ASD severity groups: LS Mean differences between Visit 8 and Visit 1. Data are presented as LS Mean (SE; P-value).**

|  | **Visit 8 – Visit 1** | | |
| --- | --- | --- | --- |
|  | Mild | Moderate | Severe |
| **Subscale 1: Language** | -6.23 (0.35; <0.0001) | -4.90 (0.26; <0.0001) | -2.91 (0.26; <0.0001) |
| **Subscale 2: Sociability** | -0.47 (0.48; 1.00) | -2.03 (0.36; <0.0001) | -3.16 (0.35; <0.0001) |
| **Subscale 3: Cognitive**  **Awareness** | interaction term not significant | interaction term not significant | interaction term not significant |
| **Subscale 4: Health** | 1.43 (0.76; <0.99) | -0.27 (0.57; 1.00) | -3.53 (0.56; <0.0001) |

**Table S4: MITA participants database: LS Mean differences between severity groups. Data are presented as: LS Mean difference (SE; P-Value)**

|  | **Visit 1** | | | **Visit 8** | | |
| --- | --- | --- | --- | --- | --- | --- |
|  | Mild vs. Moderate | Mild vs. Severe | Moderate vs. Severe | Mild vs. Moderate | Mild vs. Severe | Moderate vs. Severe |
| **Subscale 1: Language** | -0.52 (0.21; 0.87) | -0.41 (0.23; 0.99) | 0.11 (0.18; 1.00) | -1.85 (0.42; 0.006) | -3.73 (0.43; <0.0001) | -1.88 (0.36; 0.0001) |
| **Subscale 2: Sociability** | -2.53 (0.28; <0.0001) | -3.95 (0.32; <0.0001 | -1.42 (0.24; <0.0001) | 0.97 (0.57; 0.99) | -1.26 (0.59; 0.98) | -0.28 (0.47; 1.00) |
| **Subscale 3: Cognitive**  **Awareness** | interaction term not significant | interaction term not significant | interaction term not significant | interaction term not significant | interaction term not significant | interaction term not significant |
| **Subscale 4: Health** | -3.16 (0.44; <0.0001) | -5.97 (0.52; <0.0001) | -2.81 (0.39; <0.0001) | -1.46 (0.90; 1.0000) | -1.01 (0.94; 1.00) | -0.45 (0.78; 1.00) |

**Table S5: LS Means (SE; 95% CI) for Receptive Language MSEC subscale score. The differences between test and control and between Visit 12 and Visit 1 are presented as LS Mean (SE; P-value). A lower score indicates lower severity of ASD symptoms. The negative Test-Control indicates that the Test group had lower score and therefore milder symptoms.**

| **Visit Number** | **Test** | **Control** | **Test - Control** |
| --- | --- | --- | --- |
| Visit 1 | 26.7 (0.27; 26.2 - 27.2) | 27 (0.27; 26.5 - 27.5) | -0.31 (0.28; 0.2608) |
| Visit 2 | 25.1 (0.3; 24.5 - 25.7) | 26.9 (0.3; 26.3 - 27.5) | -1.75 (0.34; <0.0001) |
| Visit 3 | 24.4 (0.31; 23.7 - 25) | 25.7 (0.31; 25.1 - 26.3) | -1.34 (0.35; 0.0001) |
| Visit 4 | 23.5 (0.33; 22.9 - 24.2) | 26 (0.34; 25.3 - 26.7) | -2.47 (0.39; <0.0001) |
| Visit 5 | 23.2 (0.36; 22.5 - 23.9) | 24.7 (0.39; 24 - 25.5) | -1.54 (0.46; 0.0009) |
| Visit 6 | 22.6 (0.4; 21.9 - 23.4) | 24.6 (0.43; 23.8 - 25.5) | -2.01 (0.52; 0.0001) |
| Visit 7 | 21.9 (0.42; 21.1 - 22.7) | 24.1 (0.47; 23.2 - 25.1) | -2.24 (0.57; <0.0001) |
| Visit 8 | 21.4 (0.44; 20.6 - 22.3) | 24.5 (0.52; 23.5 - 25.5) | -3.07 (0.63; <0.0001) |
| Visit 9 | 20.4 (0.49; 19.5 - 21.4) | 23.3 (0.55; 22.3 - 24.4) | -2.9 (0.68; <0.0001) |
| Visit 10 | 20.3 (0.55; 19.2 - 21.3) | 24 (0.74; 22.5 - 25.4) | -3.7 (0.88; <0.0001) |
| Visit 11 | 19.6 (0.61; 18.4 - 20.8) | 23.4 (0.73; 22 - 24.8) | -3.85 (0.91; <0.0001) |
| Visit 12 | 18.2 (0.66; 16.9 - 19.5) | 22.1 (0.86; 20.5 - 23.8) | -3.93 (1.05; 0.0002) |
| Visit 12 – Visit 1 | -8.49 (0.65; <0.0001) | -4.87 (0.85; <0.0001) | na |

**Table S6: LS Means (SE; 95% CI) for Expressive Language measured by the Subscale 1 of ATEC. The differences between Test and Control and between Visit 12 and Visit 1 are presented as LS Mean (SE; P-value). A lower score indicates lower severity of ASD symptoms.**

| **Visit Number** | **Test** | **Control** | **Test - Control** |
| --- | --- | --- | --- |
| Visit 1 | 13.89 (0.2; 13.51 - 14.28) | 14.14 (0.19; 13.76 - 14.51) | -0.24 (0.19; 0.2141) |
| Visit 2 | 12.54 (0.22; 12.12 - 12.96) | 13.54 (0.21; 13.13 - 13.96) | -1.00 (0.23; <0.0001) |
| Visit 3 | 12.21 (0.22; 11.78 - 12.65) | 12.78 (0.22; 12.35 - 13.21) | -0.57 (0.24; 0.0194) |
| Visit 4 | 11.46 (0.23; 11.01 - 11.92) | 12.22 (0.24; 11.75 - 12.68) | -0.75 (0.27; 0.0055) |
| Visit 5 | 11.05 (0.25; 10.55 - 11.54) | 11.76 (0.27; 11.23 - 12.29) | -0.71 (0.32; 0.0241) |
| Visit 6 | 10.32 (0.27; 9.79 - 10.86) | 11.57 (0.3; 10.99 - 12.16) | -1.25 (0.36; 0.0005) |
| Visit 7 | 10.47 (0.29; 9.9 - 11.05) | 11.52 (0.32; 10.89 - 12.15) | -1.05 (0.39; 0.0071) |
| Visit 8 | 10.15 (0.31; 9.55 - 10.75) | 10.96 (0.35; 10.26 - 11.65) | -0.81 (0.43; 0.0572) |
| Visit 9 | 9.85 (0.34; 9.19 - 10.5) | 10.93 (0.37; 10.2 - 11.66) | -1.09 (0.46; 0.019) |
| Visit 10 | 9.24 (0.38; 8.51 - 9.98) | 10.77 (0.5; 9.79 - 11.76) | -1.53 (0.6; 0.0103) |
| Visit 11 | 9.16 (0.41; 8.35 - 9.97) | 10.58 (0.49; 9.62 - 11.55) | -1.42 (0.61; 0.02) |
| Visit 12 | 8.87 (0.45; 7.98 - 9.75) | 10.6 (0.58; 9.46 - 11.74) | -1.74 (0.71; 0.0144) |
| Visit 12 – Visit 1 | -5.03 (0.43; <0.0001) | -3.53 (0.57; <0.0001) | na |

**Table S7: LS Means (SE; 95% CI) for Sociability subscale score. The differences between Test and Control and between Visit 12 and Visit 1 are presented as LS Mean (SE; P-value).**

| **Visit Number** | **Test** | **Control** | **Test - Control** |
| --- | --- | --- | --- |
| Visit 1 | 13.7 (0.25; 13.2 - 14.2) | 13.8 (0.25; 13.3 - 14.3) | -0.11 (0.25; 0.6679) |
| Visit 2 | 13.1 (0.28; 12.5 - 13.6) | 13.4 (0.28; 12.8 - 13.9) | -0.32 (0.31; 0.299) |
| Visit 3 | 12.5 (0.29; 11.9 - 13) | 13.2 (0.28; 12.7 - 13.8) | -0.77 (0.32; 0.0161) |
| Visit 4 | 12.5 (0.31; 11.9 - 13.1) | 13.2 (0.31; 12.6 - 13.8) | -0.73 (0.36; 0.0424) |
| Visit 5 | 12.3 (0.33; 11.6 - 12.9) | 13.4 (0.36; 12.7 - 14.1) | -1.14 (0.42; 0.0069) |
| Visit 6 | 12.1 (0.36; 11.4 - 12.8) | 13.7 (0.4; 13 - 14.5) | -1.68 (0.48; 0.0004) |
| Visit 7 | 12 (0.39; 11.2 - 12.7) | 12.9 (0.43; 12.1 - 13.8) | -0.97 (0.52; 0.0634) |
| Visit 8 | 12.5 (0.41; 11.7 - 13.3) | 12.9 (0.48; 12 - 13.9) | -0.4 (0.58; 0.4913) |
| Visit 9 | 11.8 (0.45; 11 - 12.7) | 12.8 (0.5; 11.9 - 13.8) | -1 (0.63; 0.1101) |
| Visit 10 | 12.6 (0.5; 11.6 - 13.6) | 12.1 (0.68; 10.8 - 13.5) | 0.47 (0.81; 0.56) |
| Visit 11 | 11.8 (0.56; 10.7 - 12.9) | 13.3 (0.67; 12 - 14.6) | -1.49 (0.83; 0.0731) |
| Visit 12 | 11.8 (0.61; 10.6 - 13) | 13.1 (0.79; 11.6 - 14.7) | -1.27 (0.97; 0.1875) |
| Visit 12 – Visit 1 | -1.88 (0.59; 0.0015) | -0.72 (0.78; 0.3565) | na |

**Table S8: LS Means (SE; 95% CI) for Sensory/Cognitive Awareness subscale. The differences between Test and Control and between Visit 12 and Visit 1 are presented as LS Mean (SE; P-value).**

| **Visit Number** | **Test** | **Control** | **Test - Control** |
| --- | --- | --- | --- |
| Visit 1 | 15.4 (0.23; 15 - 15.9) | 15.4 (0.22; 15 - 15.8) | 0.02 (0.22; 0.9118) |
| Visit 2 | 14.7 (0.25; 14.2 - 15.2) | 15.3 (0.25; 14.8 - 15.7) | -0.55 (0.27; 0.042) |
| Visit 3 | 14.4 (0.26; 13.9 - 14.9) | 14.8 (0.26; 14.3 - 15.3) | -0.44 (0.28; 0.1257) |
| Visit 4 | 13.7 (0.27; 13.1 - 14.2) | 14.7 (0.28; 14.2 - 15.2) | -1.03 (0.32; 0.0013) |
| Visit 5 | 13.6 (0.3; 13 - 14.2) | 13.9 (0.32; 13.2 - 14.5) | -0.29 (0.37; 0.4434) |
| Visit 6 | 13.4 (0.32; 12.7 - 14) | 13.5 (0.35; 12.8 - 14.2) | -0.19 (0.42; 0.6587) |
| Visit 7 | 13.3 (0.34; 12.6 - 14) | 14 (0.38; 13.2 - 14.7) | -0.69 (0.46; 0.1381) |
| Visit 8 | 13.6 (0.36; 12.9 - 14.3) | 14.4 (0.42; 13.6 - 15.2) | -0.75 (0.51; 0.1393) |
| Visit 9 | 12.6 (0.4; 11.8 - 13.4) | 14.1 (0.44; 13.2 - 14.9) | -1.44 (0.55; 0.009) |
| Visit 10 | 12.8 (0.45; 11.9 - 13.6) | 14.9 (0.6; 13.7 - 16.1) | -2.17 (0.71; 0.0024) |
| Visit 11 | 12.7 (0.49; 11.8 - 13.7) | 14 (0.59; 12.9 - 15.2) | -1.29 (0.73; 0.079) |
| Visit 12 | 13 (0.54; 11.9 - 14) | 13.2 (0.7; 11.8 - 14.6) | -0.22 (0.85; 0.7987) |
| Visit 12 – Visit 1 | -2.43 (0.52; <0.0001) | -2.19 (0.68; 0.0014) | na |

**Table S9: LS Means (SE; 95% CI) for Health/Physical/Behavior subscale. The differences between Test and Control and between Visit 12 and Visit 1 are presented as LS Mean (SE; P-value).**

| **Visit Number** | **Test** | **Control** | **Test - Control** |
| --- | --- | --- | --- |
| Visit 1 | 20 (0.39; 19.3 - 20.8) | 20.8 (0.39; 20 - 21.5) | -0.73 (0.4; 0.0678) |
| Visit 2 | 19.4 (0.44; 18.5 - 20.2) | 20.5 (0.43; 19.6 - 21.3) | -1.13 (0.49; 0.0205) |
| Visit 3 | 19 (0.45; 18.1 - 19.9) | 21 (0.45; 20.1 - 21.9) | -2.02 (0.51; <0.0001) |
| Visit 4 | 19.2 (0.48; 18.2 - 20.1) | 20.5 (0.49; 19.5 - 21.5) | -1.35 (0.57; 0.0178) |
| Visit 5 | 18.5 (0.52; 17.5 - 19.5) | 20.2 (0.56; 19.1 - 21.3) | -1.71 (0.67; 0.0102) |
| Visit 6 | 17.7 (0.57; 16.6 - 18.9) | 20.9 (0.63; 19.7 - 22.1) | -3.16 (0.75; <0.0001) |
| Visit 7 | 18.4 (0.61; 17.2 - 19.6) | 21.2 (0.68; 19.9 - 22.5) | -2.81 (0.83; 0.0007) |
| Visit 8 | 18.3 (0.64; 17 - 19.6) | 21.3 (0.75; 19.8 - 22.8) | -3.02 (0.91; 0.0009) |
| Visit 9 | 17.7 (0.71; 16.4 - 19.1) | 21.3 (0.79; 19.7 - 22.8) | -3.54 (0.99; 0.0004) |
| Visit 10 | 18.5 (0.79; 16.9 - 20) | 20 (1.07; 17.9 - 22.1) | -1.52 (1.28; 0.2329) |
| Visit 11 | 18.4 (0.88; 16.6 - 20.1) | 19.7 (1.05; 17.6 - 21.8) | -1.35 (1.31; 0.3046) |
| Visit 12 | 18.4 (0.96; 16.5 - 20.2) | 19.4 (1.24; 16.9 - 21.8) | -1.02 (1.52; 0.5019) |
| Visit 12 – Visit 1 | -1.68 (0.94; 0.0726) | -1.38 (1.23; 0.2585) | na |
